# Seroresponse to SARS-CoV-2 vaccines among maintenance dialysis patients over six months

**DOI:** 10.1101/2021.09.13.21263535

**Authors:** Caroline M. Hsu, Daniel E. Weiner, Harold J. Manley, Gideon N. Aweh, Vladimir Ladik, Jill Frament, Dana Miskulin, Christos Agyropoulos, Kenneth Abreo, Andrew Chin, Reginald Gladish, Loay Salman, Doug Johnson, Eduardo K. Lacson

## Abstract

**Background and Objectives:** While most maintenance dialysis patients exhibit initial seroresponse to vaccination, concerns remain regarding the durability of this antibody response. This study evaluated immunity over time.

**Design, setting, participants, and measurements:** This retrospective cohort study included maintenance dialysis patients from a midsize national dialysis provider who received a complete SARS-CoV-2 vaccine series and had at least one antibody titer checked after full vaccination. Immunoglobulin G spike antibodies (SAb-IgG) titers were assessed monthly with routine labs beginning after full vaccination and followed over time; the semiquantitative SAb-IgG titer reported a range between 0 and ≥ 20 U/L. Descriptive analyses compared trends over time by prior history of COVID-19 and type of vaccine received. Time-to-event analyses were conducted for the outcome of loss of seroresponse (SAb-IgG < 1 U/L or development of COVID-19). Cox proportional hazards regression was used to adjust for additional clinical characteristics of interest.

**Results:** Among 1898 maintenance dialysis patients, 1567 (84%) had no prior history of COVID-19. Patients without a history of COVID-19 had declining titers over time. Among 441 BNT162b2/Pfizer recipients, median [IQR] SAb-IgG titer declined from 20 [5.99-20] U/L in month 1 to 1.30 [0.15-3.59] U/L by month 6. Among 779 mRNA-1273/Moderna recipients, median [IQR] SAb-IgG titer declined from 20 [20-20] in month 1 to 6.20 [1.74-20] by month 6. The 347 Ad26.COV2.S/Janssen recipients had a lower titer response than mRNA vaccine recipients over all time periods. In time-to-event analyses, Ad26.COV2.S/Janssen and mRNA-1273/Moderna recipients had the shortest and longest time to loss of seroresponse, respectively. The maximum titer reached in the first two months after full vaccination was predictive of the durability of the SAb-IgG seroresponse; patients with SAb-IgG titer 1-19.99 U/L were more likely to have loss of seroresponse compared to patients with SAb-IgG titer ≥ 20 U/L (HR 23.9 [95% CI: 16.1-35.5]).

**Conclusions:** Vaccine-induced seroresponse wanes over time among maintenance dialysis patients across vaccine types. Early titers after full vaccination predict the durability of seroresponse.

## Introduction

As of September 2021, the COVID-19 pandemic has claimed over 4.5 million lives worldwide, with more than 600,000 deaths in the United States.^1^ The three vaccines currently authorized for use by the Food and Drug Administration, either with approval or emergency use authorization, are all highly effective at preventing death and serious illness in the general population.^2–4^

Concerns about the robustness of the vaccine-induced immune response in vulnerable populations^5–7^ have prompted the Centers for Disease Control and Prevention (CDC) to recommend an additional dose for immunocompromised patients,^8^ while concerns about the durability of response have prompted discussion of booster doses of vaccine.^9^ The implications for patients receiving maintenance dialysis remain unclear at this time. Early studies have shown that the majority of maintenance dialysis patients generate an appropriate initial seroresponse to mRNA vaccines, although at a lower rate than the general population.^10–17^ Given that maintenance dialysis patients have an attenuated response to other vaccines, with extensive data on additional or booster doses for hepatitis B vaccination,^18^ similar concerns exist that potential uremia-associated immunocompromise may impact the response to SARS-CoV-2 vaccines. A small study in 76 dialysis patients demonstrated antibody decline in 75% of patients, with almost 20% becoming seronegative by 4 months.^19^ In the general population, waning immunity has been linked to increased breakthrough cases.^20,21^ Given the very high risk for poor outcomes associated with COVID-19 in maintenance dialysis patients, as well as their limited ability to physically distance, obtaining and maintaining immunity is of critical importance.^22,23^ We therefore conducted a retrospective multicenter study to assess the intermediate duration of vaccine-induced seroresponse among maintenance dialysis patients. Expanding on an earlier publication,^11^ we report here the seroresponse trends over time.

## Methods

Dialysis Clinic, Inc. (DCI) is a national not-for-profit provider that cares for more than 15,000 patients at 260 outpatient dialysis clinics across 29 states. Since January 2021, DCI physicians have had available an antibody monitoring protocol for patients, activated by physician order upon documentation of receipt of a SARS-CoV-2 vaccine, regardless of the vaccine type or place of administration. Like the existing hepatitis B vaccine protocol, the SARS-CoV-2 vaccine protocol documents seroresponse to vaccination by measuring antibody titers as part of the monthly blood draws. Immunoglobulin G spike antibodies (SAb-IgG) against the receptor-binding domain of the S1 subunit of SARS-CoV-2 spike antigen were measured using the chemiluminescent assay ADVIA Centaur® XP/XPT COV2G, which received emergency use authorization in July 2020.^24^ This semi-quantitative assay has a range between 0 and ≥ 20 U/L; per manufacturer specifications, SAb-IgG titer ≥ 1 U/L represents detectable antibodies, likely signifying seroresponse.^24^

Demographic and clinical data, vaccination dates, and SAb-IgG titer results were obtained from the DCI electronic health record. Patients were included if they had received a complete vaccine series of one vaccine type without additional doses. Patients were excluded from analysis if they were less than 18 years of age or did not have at least one antibody titer assessment 14 days or more after completion of a vaccine series (hereafter termed “fully vaccinated” in accordance with current CDC guidelines^25^). Baseline characteristics were assessed at the time of full vaccination. Prior COVID-19 was defined by a positive SARS-CoV-2 PCR test at any time before the date of full vaccination or SAb-IgG titer ≥ 1 U/L before or within 10 days after the first vaccine dose (representing likely prior undiagnosed COVID-19, as suggested by prior studies^2,26^). Positive SARS-CoV-2 tests were captured regardless of whether the patient was assessed in the dialysis clinic, at a testing center, or at a hospital. Analyses were stratified by prior COVID-19 status.

In analyses, SAb-IgG titers were grouped by the month of assessment relative to the date of full vaccination (month 1, 2, etc). Primary descriptive analyses compared titers by vaccine type over time. To assess whether initial vaccine response predicted sustained response, secondary descriptive analyses compared trends over time, stratifying by the maximum titer attained during the first two months following full vaccination (“maximum initial titer”). Handling of missing and duplicate values is described in the **Supplemental Methods**.

To better characterize antibody levels over time, time-to-event analysis was used to assess for the outcome of antibody titer <1 U/L or development of COVID-19, defined as having a positive SARS-CoV-2 test. Sensitivity analyses were conducted for the outcome of antibody titer <2 U/L or development of COVID-19, using a threshold suggested by DCI Lab’s internal validation methods.^11^ Of note, all DCI patients are screened for COVID-19 symptoms and recent exposure upon arrival to the dialysis facility for each treatment, followed by SARS-CoV-2 testing if they screen positive. Patients were censored at death, transplantation, or last available titer assessment. Results were compared by vaccine type and by maximum initial titer in descriptive analyses. Multivariable Cox proportional hazards regression was used to associate patients’ clinical characteristics with the outcome.

This study was reviewed and approved by the WCG IRB Work Order 1-1456342-1. Statistical analyses were performed using R v4.0.2.

## Results

Among DCI patients, 1898 adults received a full SARS-CoV-2 vaccine series and had at least one SAb-IgG titer assessment after January 1, 2021, with 1866 patients having SAb-IgG assessment following full vaccination (**Figure 1**). Of these, 1084 (58.1%) were male, 437 (23.4%) were African-American, 298 (16.0%) were Hispanic, and the average age was 63.9 ± 13.7 (SD); 299 (16.0%) had a history of COVID-19 based on either early positive SAb-IgG levels or prior positive testing. BNT162b2/Pfizer recipients tended to be older and have longer follow-up, while Ad26.COV2.S/Janssen recipients tended to be of Black race or non-Hispanic ethnicity. Among the 1567 patients without a history of COVID-19, a higher proportion of mRNA-1273/Moderna recipients were receiving peritoneal dialysis compared to BNT162b2/Pfizer and Ad26.COV2.S/Janssen recipients (**Table 1**).

**Figure 1.**
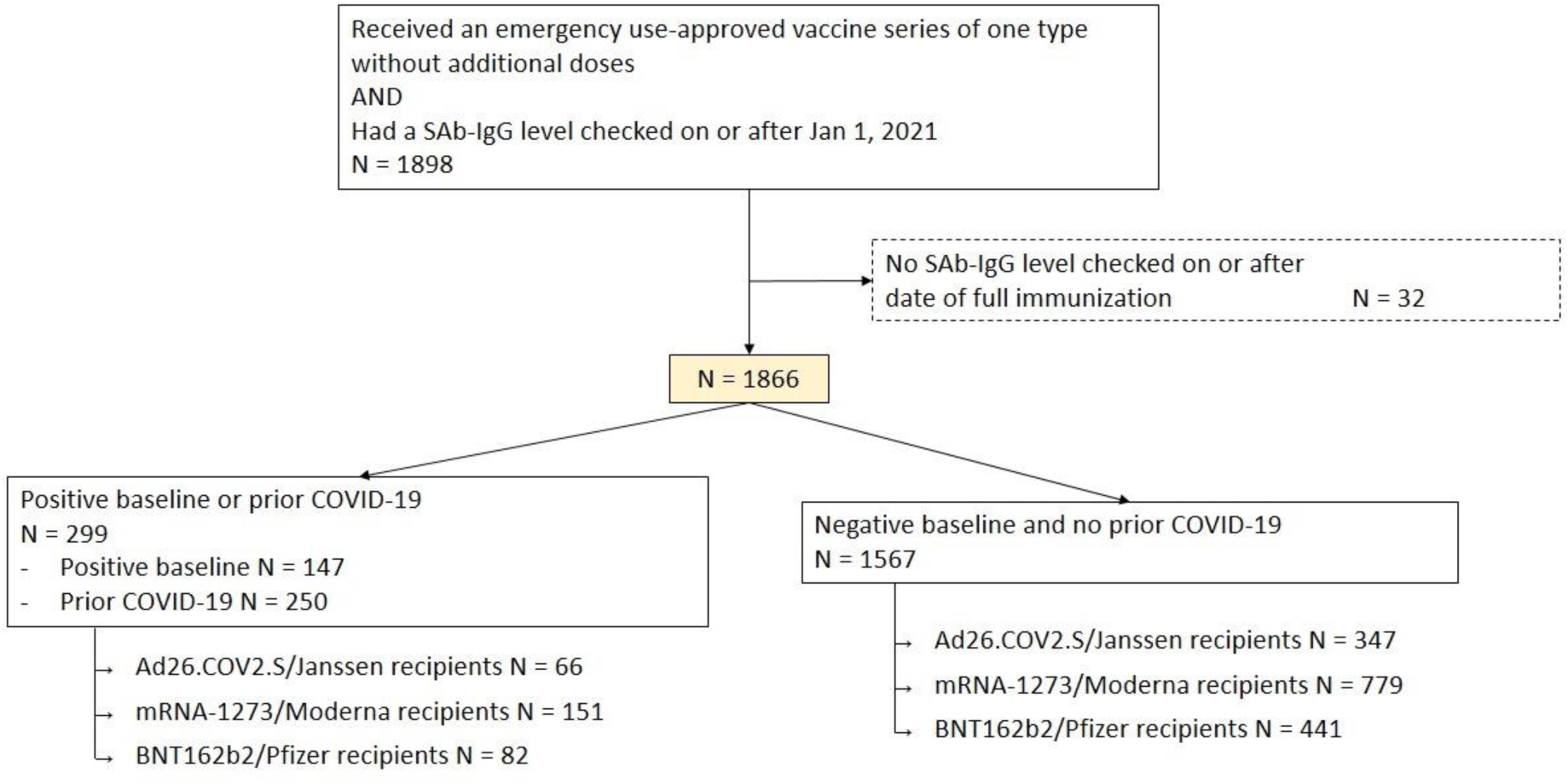
Flow diagram. Baseline defined by SAb-IgG titer > 1 before or within 10 days after first dose of vaccine Prior COVID-19 defined as positive SARS-CoV-2 test before full immunity (at 14 days after completion of a vaccine series)

**Table 1.** Baseline patient characteristics by history of COVID-19 and type of vaccine received.

|  |  | No history of COVID-19 (N=1567) |  |  | History of COVID-19 (N=299) |  |  |
| --- | --- | --- | --- | --- | --- | --- | --- |
|  | Overall | Ad26.COV2.S/<br>Janssen | mRNA-1273/<br>Moderna | BNT162b2/<br>Pfizer | Ad26.COV2.S/<br>Janssen | mRNA-1273/<br>Moderna | BNT162b2/<br>Pfizer |
| n | 1866 | 347 | 779 | 441 | 66 | 151 | 82 |
| Age | 63.9 ± 13.7 | 61.8 ± 12.6 | 63.7 ± 14.3 | 67.0 ± 13.5 | 65.1 ± 12.6 | 59.8 ± 12.0 | 64.2 ± 13.6 |
| Male sex | 1084 (58.1) | 189 (54.5) | 471 (60.5) | 254 (57.6) | 36 (54.5) | 82 (54.3) | 52 (63.4) |
| Race |  |  |  |  |  |  |  |
| Native American | 145 (7.8) | 6 (1.7) | 50 (6.4) | 55 (12.5) | 1 (1.5) | 21 (13.9) | 12 (14.6) |
| Asian | 105 (5.6) | 7 (2.0) | 35 (4.5) | 49 (11.1) | 0 (0.0) | 6 (4.0) | 8 (9.8) |
| Black | 437 (23.4) | 169 (48.7) | 118 (15.1) | 81 (18.4) | 31 (47.0) | 20 (13.2) | 18 (22.0) |
| Unknown/Other | 225 (12.1) | 34 (9.8) | 114 (14.6) | 47 (10.7) | 6 (9.1) | 18 (11.9) | 6 (7.3) |
| White | 954 (51.1) | 131 (37.8) | 462 (59.3) | 209 (47.4) | 28 (42.4) | 86 (57.0) | 38 (46.3) |
| Hispanic ethnicity | 298 (16.0) | 10 (2.9) | 148 (19.0) | 60 (13.6) | 8 (12.1) | 59 (39.1) | 13 (15.9) |
| Vintage, months | 36.6 [15.5, 70.1] | 38.2 [13.8, 81.9] | 36.1 [16.2, 69.4] | 34.3 [14.7, 62.2] | 36.8 [17.3, 84.1] | 39.4 [17.8, 60.8] | 40.4 [17.9, 87.3] |
| Body mass index, kg/m<sup>2</sup> | 28.5 ± 7.2 | 29.3 ± 8.0 | 28.2 ± 6.6 | 28.1 ± 7.3 | 29.6 ± 7.4 | 28.4 ± 8.1 | 28.0 ± 6.5 |
| Diabetes | 1097 (58.8) | 215 (62.0) | 436 (56.0) | 252 (57.1) | 42 (63.6) | 98 (64.9) | 54 (65.9) |
| Long-term care facility | 85 (4.6) | 9 (2.6) | 26 (3.3) | 19 (4.3) | 5 (7.6) | 10 (6.6) | 16 (19.5) |
| Modality |  |  |  |  |  |  |  |
| Home hemodialysis | 38 (2.0) | 1 (0.3) | 26 (3.3) | 10 (2.3) | 0 (0.0) | 1 (0.7) | 0 (0.0) |
| In-center hemodialysis | 1603 (85.9) | 311 (89.6) | 634 (81.4) | 380 (86.2) | 60 (90.9) | 139 (92.1) | 79 (96.3) |
| Peritoneal dialysis | 225 (12.1) | 35 (10.1) | 119 (15.3) | 51 (11.6) | 6 (9.1) | 11 (7.3) | 3 (3.7) |
| Inadequate dialysis | 55 (2.9) | 17 (4.9) | 16 (2.1) | 11 (2.5) | 2 (3.0) | 6 (4.0) | 3 (3.7) |
| Albumin, g/dL | 3.9 ± 0.4 | 3.9 ± 0.4 | 3.9 ± 0.4 | 3.9 ± 0.5 | 3.9 ± 0.4 | 3.9 ± 0.4 | 3.8 ± 0.4 |
| HBsAb ≥10 mIU/mL | 1431 (76.7) | 251 (72.3) | 606 (77.8) | 331 (75.1) | 49 (74.2) | 132 (87.4) | 62 (75.6) |
| History of transplantation | 107 (5.7) | 17 (4.9) | 46 (5.9) | 28 (6.3) | 4 (6.1) | 5 (3.3) | 7 (8.5) |
| Immunodeficiency | 83 (4.4) | 18 (5.2) | 33 (4.2) | 23 (5.2) | 3 (4.5) | 5 (3.3) | 1 (1.2) |
| Immunomodulating medication | 242 (13.0) | 43 (12.4) | 93 (11.9) | 70 (15.9) | 9 (13.6) | 18 (11.9) | 9 (11.0) |
| Duration of follow-up, days | 112 [91, 134] | 110 [104, 118] | 116 [93, 130] | 121 [92, 155] | 113 [107, 135] | 98 [81, 125] | 132 [89, 157] |

Vintage and duration of follow-up are reported as median [IQR]. All other data are reported as mean ± standard deviation or %.
Inadequate dialysis defined by hemodialysis dose spKt/V<1.2 or peritoneal dialysis dose weekly Kt/V<1.7
HBsAb ≥10 mIU/mL signifies hepatitis B seroimmunity

Patients without a history of COVID-19 who received mRNA vaccine had declining SAb-IgG titers over time (**Figure 2A**). Among BNT162b2/Pfizer recipients, median [IQR, N number of data values] antibody titer was 20 [5.99-20, N of 339] U/L in month 1, with a reduction to 2.69 [0.70-9.38, N of 337] by month 4 and 1.30 [0.15-3.59, N of 174] U/L by month 6. Among mRNA-1273/Moderna recipients, median [IQR, N number of data values] SAb-IgG titer declined from 20 [20-20, N of 646] in month 1 to 20 [4.03-20, N of 604] by month 4 and to 6.20 [1.74-20, N of 85] by month 6. Over all time periods, Ad26.COV2.S/Janssen recipients had median [IQR] SAb-IgG titer of less than 1 U/L, without significant change over time. Among patients with a history of COVID-19, more than 75% of mRNA vaccine recipients maintained antibody titer at the upper limit of the assay’s detection (20 U/L) through month 6; this is true also of Janssen vaccine recipients through month 5 (with 6-month data limited by the number of participants with follow-up to this time point) (**Figure 2B**).

**Figure 2.**
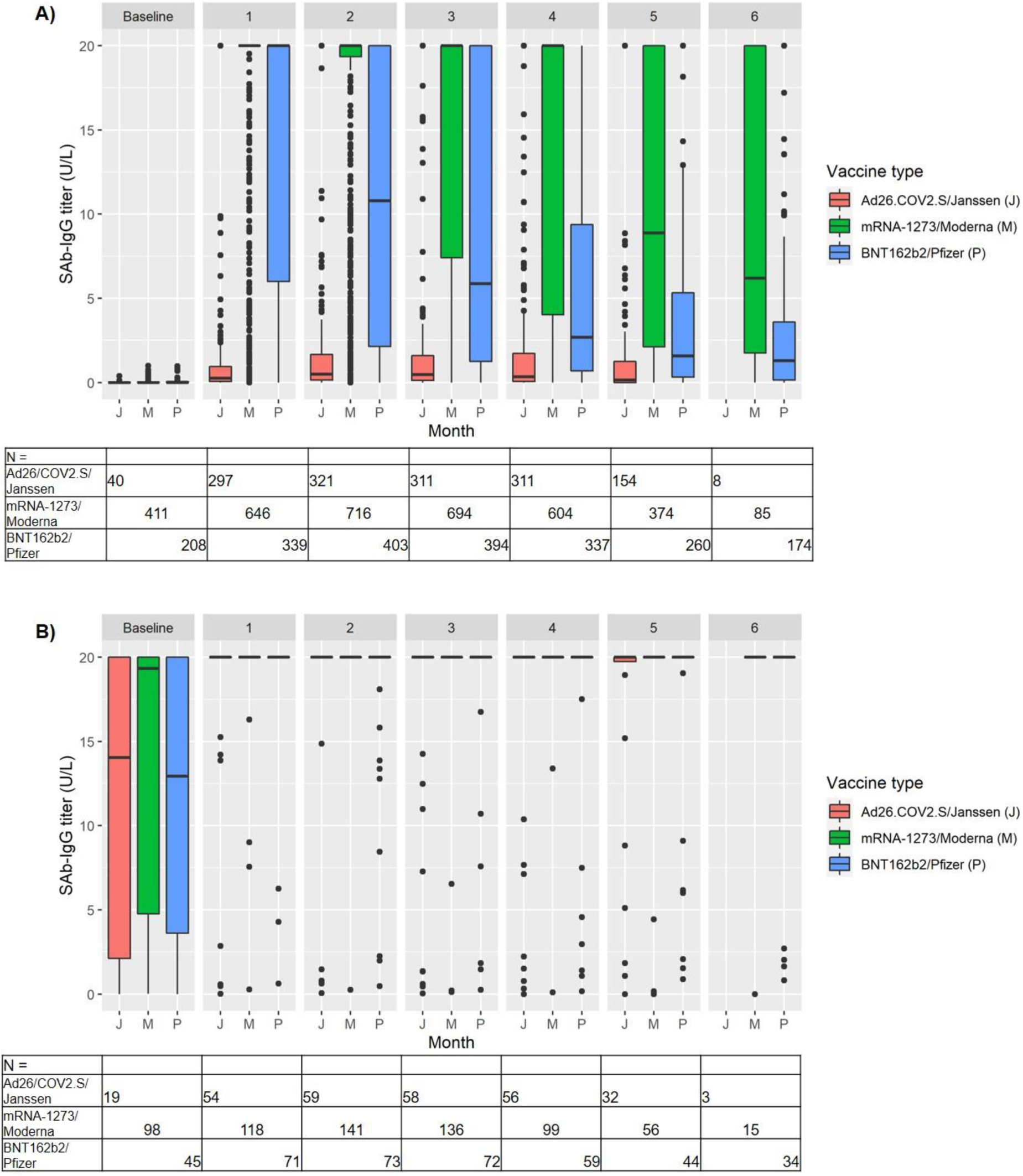
SAb-IgG titers vs months after date of full immunization, comparing by vaccine type. A) Patients without prior COVID-19 B) Patients with prior COVID-19 The tables of N show the number of titers for each month, by vaccine type. Data are not shown for groups with N < 10

Among the 1567 patients without a history of COVID-19, 559 developed SAb-IgG titer < 1 U/L or were diagnosed with COVID-19 (556 and 3 patients, respectively). Time-to-event analysis showed a difference by vaccine type, with Ad26COV2.S/Janssen recipients having the shortest time to SAb-IgG < 1 U/L while mRNA-1273/Moderna recipients had more durable SAb-IgG titer levels (**Figure 3**). At month 4, 67.5% of Janssen, 32.1% of Pfizer and 12.3% of Moderna recipients had SAb-IgG titers less than 1 U/L; at month 6, 43.9% of Pfizer and 14.1% of Moderna recipients had SAb-IgG titers less than 1 U/L (as noted, 6-month data on Janssen recipients is limited by the number of participants with follow-up to this time point). Sensitivity analysis using the threshold of SAb-IgG titer <2 instead showed more events occurring, as expected, but the differences by vaccine type remained (**Supplemental Figure S1**).

**Figure 3.**
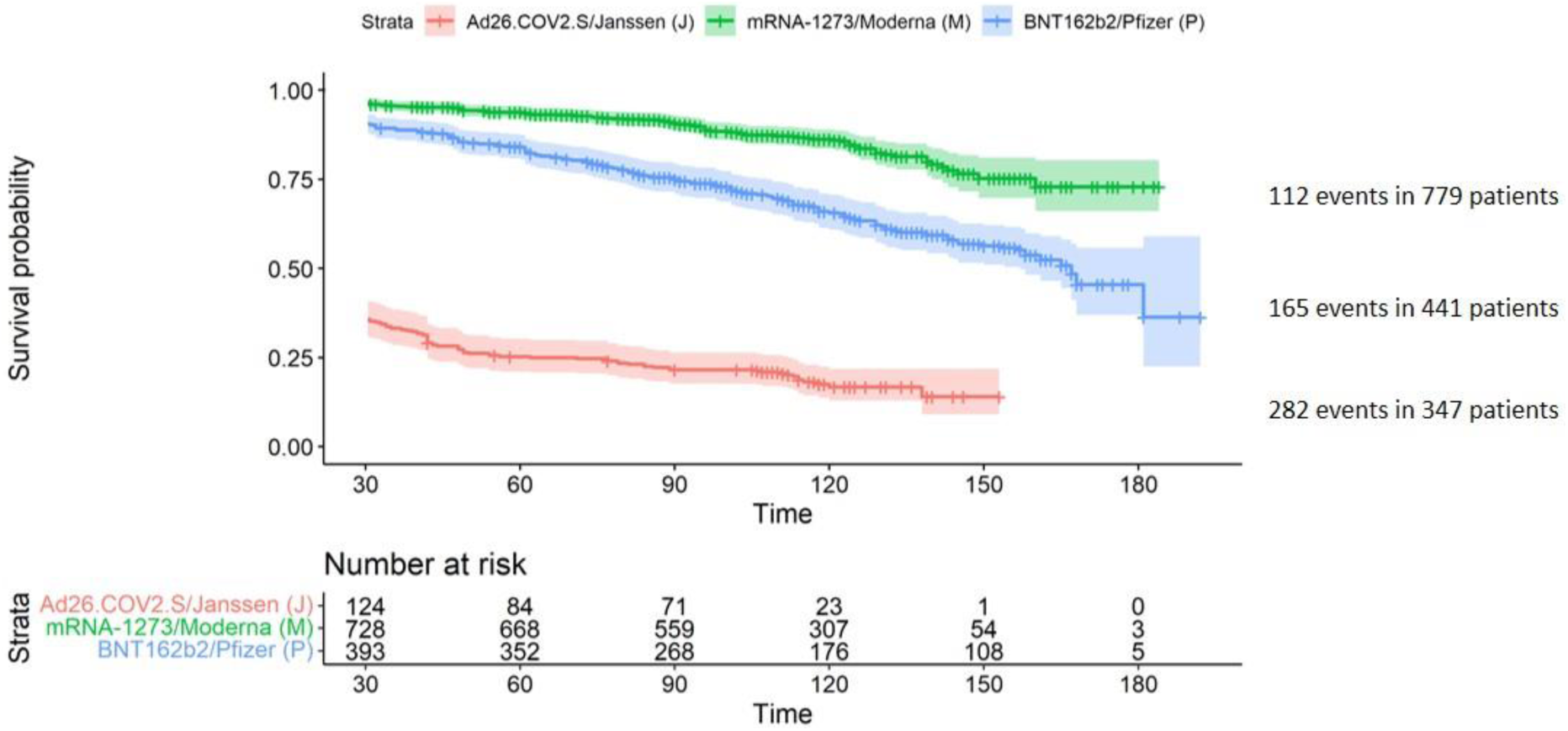
Kaplan-Meier survival curves for the outcome of Ab titer <1 or diagnosis of COVID-19, among those without a history of COVID-19, by vaccine type. Data are shown beginning at Day 30, at which time all patients have had at least one opportunity for assessment of the outcome of Ab titer < 1 U/L via monthly labs. Patients were censored at death, transplantation, or last available titer assessment.

To assess whether initial seroresponse predicts sustained response, patients were then grouped by maximum titer measured during the first two months following full vaccination: 866, 345, and 302 patients had maximum initial titer of ≥ 20, 1 to 19.99, and less than 1 U/L, respectively, while 54 patients did not have a titer assessed during the first two months of full immunity and were excluded from this analysis. Black race, non-Hispanic ethnicity, and use of immunomodulating medications were more likely to be associated with a maximum initial titer of less than 1. Moderna recipients were more likely to have a maximum initial titer of 20 U/L or higher, and Janssen recipients were more likely to have a maximum initial titer of less than 1 U/L (**Table 2**).

**Table 2.** Baseline patient characteristics by maximum initial titer, among those without a history of COVID-19.

|  | SAb-IgG ≥20 U/L | SAb-IgG 1-19.99 U/L | SAb-IgG <1 U/L |
| --- | --- | --- | --- |
| n | 866 | 345 | 302 |
| Age | 62.4 ± 13.9 | 66.9 ± 13.5 | 65.5 ± 12.8 |
| Male sex | 531 (61.3) | 187 (54.2) | 166 (55.0) |
| Race |  |  |  |
| Native American | 74 (8.5) | 30 (8.7) | 4 (1.3) |
| Asian/Pacific Islander | 62 (7.2) | 18 (5.2) | 11 (3.6) |
| Black | 176 (20.3) | 81 (23.5) | 105 (34.8) |
| Unknown/Other | 115 (13.3) | 34 (9.9) | 35 (11.6) |
| White | 439 (50.7) | 182 (52.8) | 147 (48.7) |
| Hispanic ethnicity | 157 (18.1) | 39 (11.3) | 19 (6.3) |
| Vintage, months | 36.9 [16.4, 72.1] | 37.0 [14.0, 67.8] | 33.4 [13.5, 70.4] |
| Body mass index, kg/m<sup>2</sup> | 28.4 ± 6.9 | 27.8 ± 6.9 | 29.2 ± 7.9 |
| Diabetes | 489 (56.5) | 199 (57.7) | 184 (60.9) |
| Long-term care facility | 27 (3.1) | 11 (3.2) | 13 (4.3) |
| Modality |  |  |  |
| Home hemodialysis | 25 (2.9) | 6 (1.7) | 0 (0.0) |
| In-center hemodialysis | 736 (85.0) | 290 (84.1) | 263 (87.1) |
| Peritoneal dialysis | 105 (12.1) | 49 (14.2) | 39 (12.9) |
| Inadequate dialysis | 20 (2.3) | 9 (2.6) | 13 (4.3) |
| Albumin, g/dL | 3.9 ± 0.4 | 3.8 ± 0.5 | 3.8 ± 0.4 |
| HBsAb ≥10 mIU/mL | 707 (81.6) | 256 (74.2) | 193 (63.9) |
| History of transplantation | 49 (5.7) | 15 (4.3) | 26 (8.6) |
| Immunodeficiency | 37 (4.3) | 16 (4.6) | 20 (6.6) |
| Immunomodulating medication | 92 (10.6) | 42 (12.2) | 67 (22.2) |
| Vaccine type |  |  |  |
| Ad26.COV2.S/Janssen | 36 (4.2) | 87 (25.2) | 217 (71.9) |
| mRNA-1273/Moderna | 595 (68.7) | 127 (36.8) | 30 (9.9) |
| BNT162b2/Pfizer | 235 (27.1) | 131 (38.0) | 55 (18.2) |
| Duration of follow-up, days | 116 [94, 137] | 118 [101, 141] | 105 [83, 118] |

Vintage and duration of follow-up are reported as median [IQR]. All other data are reported as mean ± standard deviation or %.
Inadequate dialysis defined by hemodialysis dose spKt/V<1.2 or peritoneal dialysis dose weekly Kt/V<1.7
HBsAb ≥10 mIU/mL signifies hepatitis B seroimmunity

In time-to-event analysis, those who had a maximum initial titer ≥ 20 U/L were less likely to develop the outcome of titer less than 1 U/L or COVID-19 compared to those with a maximum initial titer from 1 to 19.99 U/L. This difference persisted even among recipients of the same vaccine type (**Figures 4A-D**) and in sensitivity analyses for the outcome of titer less than 2 U/L (with corresponding change in strata thresholds, **Supplemental Figures S2A-D**). In multivariate Cox proportional hazards regression, in addition to differences by vaccine type, older age, White race, higher body mass index, lower albumin, lack of hepatitis B seroimmunity, and use of immunomodulating medications were associated with shorter time to loss of seroresponse (**Figure 5**). In sensitivity analyses, body mass index was no longer associated with loss of seroresponse; other findings were similar (**Supplemental Figure S3)**.

**Figure 4.**
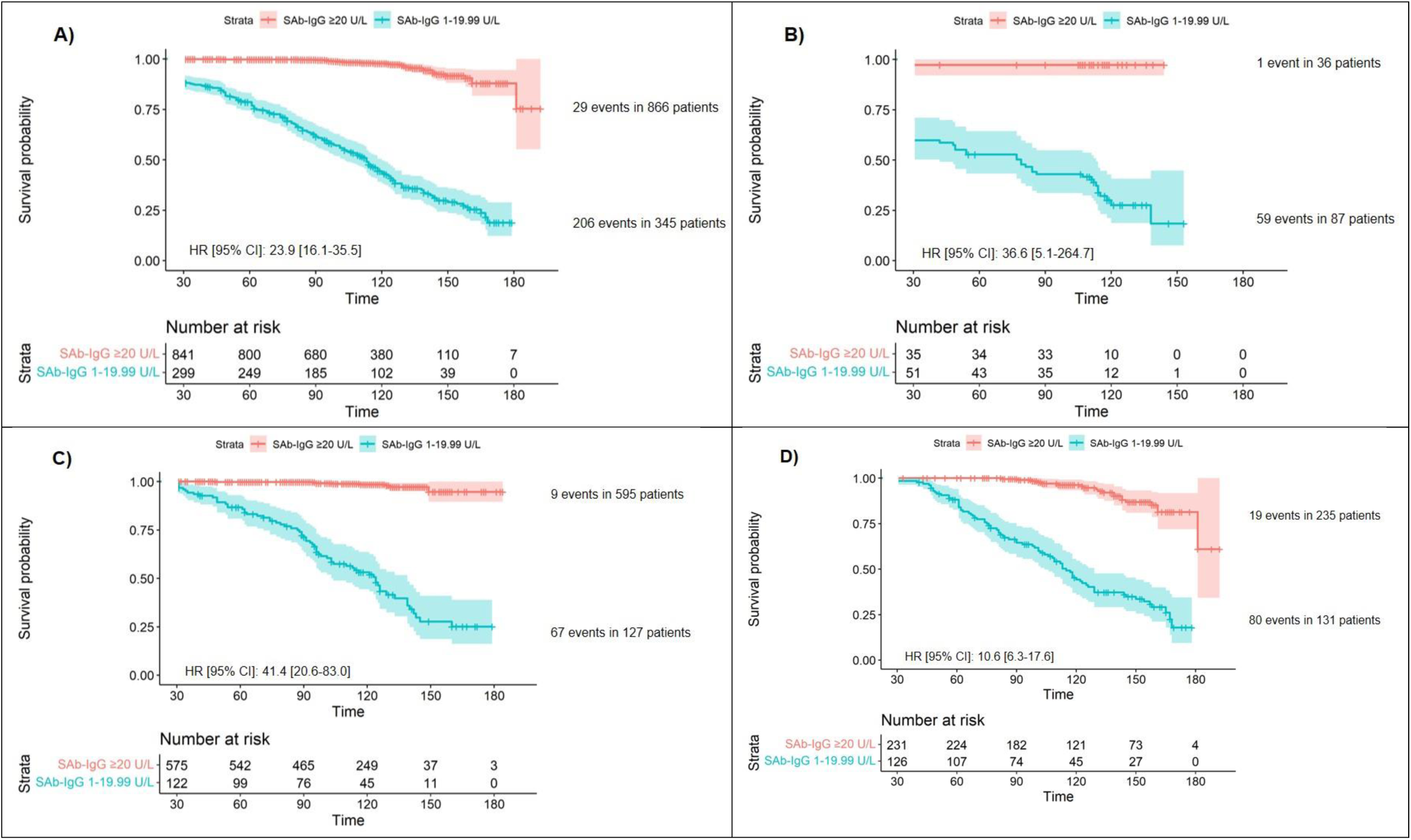
Kaplan-Meier survival curves for the outcome of SAb-IgG titer <1 or diagnosis of COVID-19, among those without a history of COVID-19, by maximum initial SAb-IgG titer. Data are shown beginning at Day 30, at which time all patients have had at least one opportunity for assessment of the outcome of SAb-IgG titer < 1 U/L via monthly labs. Patients were censored at death, transplantation, or last available titer assessment. Patients with maximum initial SAb-IgG titer are not shown since, given our definition of maximum initial titer, all had experienced the outcome by the end of month 2. A) All patients B) Ad26.COV2.S/Janssen recipients only C) mRNA-1273/Moderna recipients only D) BNT162b2/Pfizer recipients only

**Figure 5.**
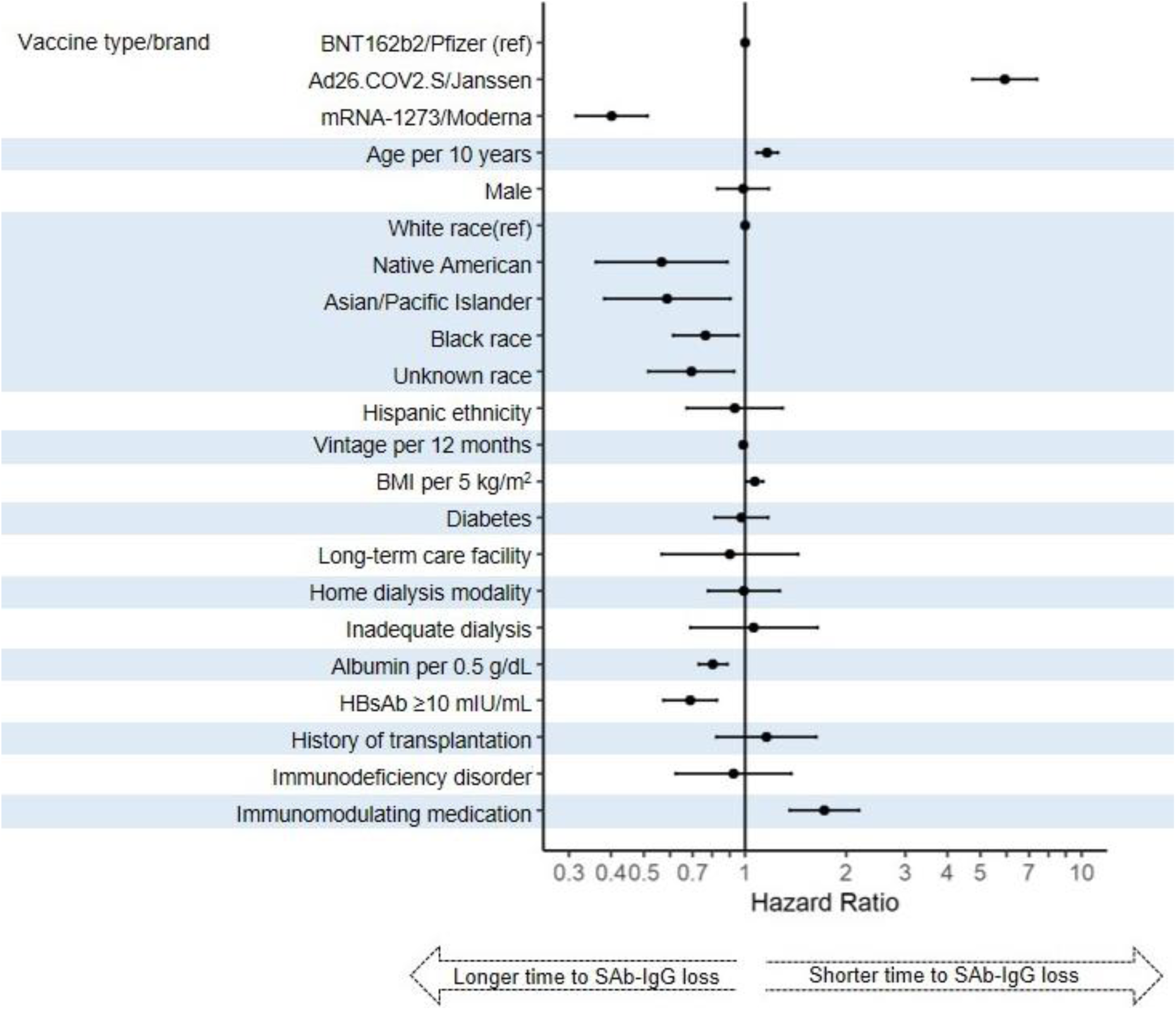
Multivariable Cox proportional hazards regression of clinical characteristics predicting loss of SAb-IgG seroresponse (outcome of SAb-IgG titer < 1 or development of COVID-19). Inadequate dialysis defined by hemodialysis dose spKt/V<1.2 or peritoneal dialysis dose weekly Kt/V<1.7 HBsAb ≥10 mIU/mL signifies hepatitis B seroimmunity

## Discussion

Among a national population of maintenance dialysis patients in the United States, mRNA vaccines elicited greater seroresponse than the Ad26.COV2.S/Janssen vaccine, but antibody titers against the SARS-CoV-2 spike protein waned substantially over the first six months following full vaccination among patients without a prior history of COVID-19. Furthermore, the robustness of the initial antibody response appears to predict the rapidity of subsequent waning of antibody levels. Other predictors of the duration of seroresponse largely reflect patients’ immune health.

As of August 13, 2021, the CDC recommends a third dose of mRNA vaccine for those with moderate to severe immunocompromise.^8^ The inclusion of maintenance dialysis patients within this classification is non-specific and depends upon the medical judgment of the treating clinician. Critically, initial studies suggest that vaccine-induced immunity among maintenance dialysis patients immediately following receipt of a vaccine series was intermediate, with somewhat lesser response than among the general population but a greater response than among patients receiving immunosuppression for transplantation or other indications.^10–17^ Our data provide longitudinal evidence of immunity waning fairly rapidly over time, contrasting somewhat with SAb-IgG titers measured over time in healthy adults.^27–30^ These data estimate that, even following current vaccination standards, about half of maintenance dialysis patients will have suboptimal vaccine-induced protection in the fall of 2021, a time of projected high community prevalence of COVID-19.^31,32^ While the antibody level needed for protection from disease has not been definitively determined, a general correlation between seroimmunity and protection from disease has been observed.^33^ Additionally, individuals with seroimmunity may be less likely to transmit the virus, providing protection to other vulnerable patients in mandatory congregate healthcare settings.^34,35^ Similar data on waning immunity prompted officials in France to recommend an additional vaccine dose to maintenance dialysis patients in April 2021, with some early evidence of subsequent strengthened immunity in several small studies.^26,36–38^

There may be a role for wider routine monitoring of SAb-IgG titers for maintenance dialysis patients. Seroimmunity to hepatitis B is currently monitored through such a protocol, and, due to this population’s regular and frequent contact with the medical system, routine testing and administration of additional doses would not be logistically difficult. In particular, the peak response in the first two months of full immunity indicates one’s likely course and could be used to anticipate the timing of loss of seroimmunity.^30^ Of note, the CDC currently does not recommend using COVID-19 antibody testing to guide clinical decision-making.^39^

This study represents a real-world, geographically diverse, multi-center population of maintenance dialysis patients in the United States, in whom prior risk of COVID-19 morbidity and mortality has been documented.^23,40^ We acknowledge this study’s limitations. As with all observational studies, confounding variables may affect interpretation of results. In particular, data for months 5 and 6 primarily reflect patients who received their doses vaccine early (e.g. January and February of 2021) with a moderately higher proportion of BNT162b2/Pfizer vaccine recipients, and these patients may be frailer at baseline. In addition, we did not correlate antibody titers with breakthrough infection, an issue which remains complex and controversial.

In conclusion, immunity to SARS-CoV-2 vaccines, as indicated by SAb-IgG titers, wanes over time in maintenance dialysis patients without a prior history of COVID-19. In the setting of SARS-CoV-2 variants of concern, the impact of waning titers on breakthrough infections needs to be monitored closely. The current CDC recommendation to provide a third vaccine dose based on clinical assessment for immune compromise is an important consideration for the maintenance dialysis population. A large proportion of maintenance dialysis patients have suboptimal response to the currently recommended vaccine regimens. Therefore, additional doses of vaccine should be considered for this vulnerable population, whether routinely or, with further investigation, potentially guided by protective correlates such as antibody response.

## Data Availability

Data are available upon request.

## References

1. WHO Coronavirus Disease (COVID-19) Dashboard. World Health Organization; 2021. Accessed September 6, 2021. https://covid19.who.int/

2. Polack FP, Thomas SJ, Kitchin N, et al. Safety and Efficacy of the BNT162b2 mRNA Covid-19 Vaccine. N Engl J Med. 2020;383(27):2603–2615. doi:10.1056/NEJMoa2034577

3. Baden LR, El Sahly HM, Essink B, et al. Efficacy and Safety of the mRNA-1273 SARS-CoV-2 Vaccine. N Engl J Med. 2021;384(5):403–416. doi:10.1056/NEJMoa2035389

4. Sadoff J, Gray G, Vandebosch A, et al. Safety and Efficacy of Single-Dose Ad26.COV2.S Vaccine against Covid-19. N Engl J Med. 2021;384(23):2187–2201. doi:10.1056/NEJMoa2101544

5. Tenforde MW, Patel MM, Ginde AA, et al. Effectiveness of SARS-CoV-2 MRNA Vaccines for Preventing Covid-19 Hospitalizations in the United States. Infectious Diseases (except HIV/AIDS); 2021. doi:10.1101/2021.07.08.21259776

6. Brosh-Nissimov T, Orenbuch-Harroch E, Chowers M, et al. BNT162b2 vaccine breakthrough: clinical characteristics of 152 fully vaccinated hospitalized COVID-19 patients in Israel. Clinical Microbiology and Infection. Published online July 2021:S1198743×21003670. doi:10.1016/j.cmi.2021.06.036

7. Chemaitelly H, AlMukdad S, Joy JP, et al. SARS-CoV-2 Vaccine Effectiveness in Immunosuppressed Kidney Transplant Recipients. Epidemiology; 2021. doi:10.1101/2021.08.07.21261578

8. Media Statement from CDC Director Rochelle P. Walensky, MD, MPH, on Signing the Advisory Committee on Immunization Practices’ Recommendation for an Additional Dose of an MRNA COVID-19 Vaccine in Moderately to Severely Immunocompromised People. Centers for Disease Control and Prevention; 2021. https://www.cdc.gov/media/releases/2021/s0813-additional-mRNA-mrna-dose.html

9. Joint Statement from HHS Public Helath and Medical Experts on COVID-19 Booster Shots.; 2021. Accessed September 1, 2021. https://www.hhs.gov/about/news/2021/08/18/joint-statement-hhs-public-health-and-medical-experts-covid-19-booster-shots.html

10. Carr EJ, Kronbichler A, Graham-Brown M, et al. Systematic Review of Early Immune Response to SARS-CoV-2 Vaccination Among Patients with Chronic Kidney Disease. Kidney Int Rep. Published online July 6, 2021. doi:10.1016/j.ekir.2021.06.027

11. Lacson E, Argyropoulos C, Manley H, et al. Immunogenicity of SARS-CoV-2 Vaccine in Dialysis. JASN. Published online August 4, 2021:ASN.2021040432. doi:10.1681/ASN.2021040432

12. Longlune N, Nogier MB, Miedougé M, et al. High immunogenicity of a messenger RNA-based vaccine against SARS-CoV-2 in chronic dialysis patients. Nephrology Dialysis Transplantation. Published online May 31, 2021:gfab193. doi:10.1093/ndt/gfab193

13. Anand S, Montez-Rath M, Han J, et al. Antibody Response to COVID-19 Vaccination in Patients Receiving Dialysis. J Am Soc Nephrol. Published online June 11, 2021. doi:10.1681/ASN.2021050611

14. Agur T, Ben-Dor N, Goldman S, et al. Antibody response to mRNA SARS-CoV-2 vaccine among dialysis patients - a prospectivecohort study. Nephrol Dial Transplant. Published online April 11, 2021. doi:10.1093/ndt/gfab155

15. Chan L, Fuca N, Zeldis E, Campbell KN, Shaikh A. Antibody Response to mRNA-1273 SARS-CoV-2 Vaccine in Hemodialysis Patients with and without Prior COVID-19. CJASN. Published online May 24, 2021:CJN.04080321. doi:10.2215/CJN.04080321

16. Labriola L, Scohy A, Van Regemorter E, et al. Immunogenicity of BNT162b2 SARS-CoV-2 Vaccine in a Multicenter Cohort of Nursing Home Residents Receiving Maintenance Hemodialysis. American Journal of Kidney Diseases. Published online August 2021:S0272638621007745. doi:10.1053/j.ajkd.2021.07.004

17. Danthu C, Hantz S, Dahlem A, et al. Humoral Response after SARS-Cov-2 mRNA Vaccine in a Cohort of Hemodialysis Patients and Kidney Transplant Recipients. J Am Soc Nephrol. Published online June 16, 2021:ASN.2021040490. doi:10.1681/ASN.2021040490

18. Chi C, Patel P, Pilishvili T, Moore M, Murphy T, Strikas R. Guidelines for Vaccinating Kidney Dialysis Patients and Patients with Chronic Kidney Disease. Centers for Disease Control and Prevention; 2012. https://www.cdc.gov/dialysis/pdfs/vaccinating_dialysis_patients_and_patients_dec2012.pdf

19. Dulovic A, Strengert M, Ramos GM, et al. Diminishing Immune Responses against Variants of Concern in Dialysis Patients Four Months after SARS-CoV-2 MRNA Vaccination. Nephrology; 2021. doi:10.1101/2021.08.16.21262115

20. Mizrahi B, Lotan R, Kalkstein N, et al. Correlation of SARS-CoV-2 Breakthrough Infections to Time-from-Vaccine; Preliminary Study. Epidemiology; 2021. doi:10.1101/2021.07.29.21261317

21. Bergwerk M, Gonen T, Lustig Y, et al. Covid-19 Breakthrough Infections in Vaccinated Health Care Workers. N Engl J Med. Published online July 28, 2021:NEJMoa2109072. doi:10.1056/NEJMoa2109072

22. Chung EY, Palmer SC, Natale P, et al. Incidence and Outcomes of COVID-19 in People With CKD: A Systematic Review and Meta-analysis. Am J Kidney Dis. Published online August 5, 2021:S0272-6386(21)00771-X. doi:10.1053/j.ajkd.2021.07.003

23. Hsu CM, Weiner DE, Aweh G, et al. COVID-19 Among US Dialysis Patients: Risk Factors and Outcomes From a National Dialysis Provider. Am J Kidney Dis. 2021;77(5):748-756.e1. doi:10.1053/j.ajkd.2021.01.003

24. COV2G, ADVIA Centaur XP and ADVIA Centaur CPT Systems. Published online July 2020. Accessed June 22, 2021. https://www.fda.gov/media/140704/download

25. COVID-19: When You’ve Been Fully Vaccinated. Centers for Disease Control and Prevention; 2021. https://www.cdc.gov/coronavirus/2019-ncov/vaccines/fully-vaccinated.html

26. Espi M, Charmetant X, Barba T, et al. Justification, Safety, and Efficacy of a Third Dose of MRNA Vaccine in Maintenance Hemodialysis Patients: A Prospective Observational Study. Nephrology; 2021. doi:10.1101/2021.07.02.21259913

27. Doria-Rose N, Suthar MS, Makowski M, et al. Antibody Persistence through 6 Months after the Second Dose of mRNA-1273 Vaccine for Covid-19. N Engl J Med. 2021;384(23):2259–2261. doi:10.1056/NEJMc2103916

28. Pegu A, O’Connell S, Schmidt SD, et al. Durability of MRNA-1273-Induced Antibodies against SARS-CoV-2 Variants. Immunology; 2021. doi:10.1101/2021.05.13.444010

29. Barouch DH, Stephenson K, Sadoff J, et al. Durable Humoral and Cellular Immune Responses Following Ad26.COV2.S Vaccination for COVID-19. Infectious Diseases (except HIV/AIDS); 2021. doi:10.1101/2021.07.05.21259918

30. Favresse J, Bayart J-L, Mullier F, et al. Antibody titres decline 3-month post-vaccination with BNT162b2. Emerg Microbes Infect. 2021;10(1):1495–1498. doi:10.1080/22221751.2021.1953403

31. COVID-19 Forecasts: Cases. Centers for Disease Control and Prevention; 2021. Accessed September 6, 2021. https://www.cdc.gov/coronavirus/2019-ncov/science/forecasting/forecasts-cases.html

32. COVID-19 Projections. Institute for Health Metrics and Evaluation; 2021. Accessed September 6, 2021. https://covid19.healthdata.org/united-states-of-america?view=daily-deaths&tab=trend

33. Earle KA, Ambrosino DM, Fiore-Gartland A, et al. Evidence for antibody as a protective correlate for COVID-19 vaccines. Vaccine. 2021;39(32):4423–4428. doi:10.1016/j.vaccine.2021.05.063

34. Harris RJ, Hall JA, Zaidi A, Andrews NJ, Dunbar JK, Dabrera G. Effect of Vaccination on Household Transmission of SARS-CoV-2 in England. N Engl J Med. 2021;385(8):759–760. doi:10.1056/NEJMc2107717

35. Prunas O, Warren JL, Crawford FW, et al. Vaccination with BNT162b2 Reduces Transmission of SARS-CoV-2 to Household Contacts in Israel. Infectious Diseases (except HIV/AIDS); 2021. doi:10.1101/2021.07.13.21260393

36. Dekervel M, Henry N, Torreggiani M, et al. Humoral response to a third injection of BNT162b2 vaccine in patients on maintenance hemodialysis. Clinical Kidney Journal. Published online August 13, 2021:sfab152. doi:10.1093/ckj/sfab152

37. Ducloux D, Colladant M, Chabannes M, Yannaraki M, Courivaud C. Humoral response after 3 doses of the BNT162b2 mRNA COVID-19 vaccine in patients on hemodialysis. Kidney International. 2021;100(3):702–704. doi:10.1016/j.kint.2021.06.025

38. Bensouna I, Caudwell V, Kubab S, et al. SARS-CoV-2 Antibody Response After a Third Dose of the BNT162b2 Vaccine in Patients Receiving Maintenance Hemodialysis or Peritoneal Dialysis. American Journal of Kidney Diseases. Published online September 2021:S0272638621008337. doi:10.1053/j.ajkd.2021.08.005

39. Summary Document for Interim Clinical Considerations for Use of COVID-19 Vaccines Currently Authorized in the United States. Centers for Disease Control and Prevention; 2021. Accessed August 31, 2021. https://www.cdc.gov/vaccines/covid-19/downloads/summary-interim-clinical-considerations.pdf

40. Hsu CM, Weiner DE, Aweh G, Salenger P, Johnson DS, Lacson E. Epidemiology and Outcomes of COVID-19 in Home Dialysis Patients Compared with In-Center Dialysis Patients. J Am Soc Nephrol. 2021;32(7):1569–1573. doi:10.1681/ASN.2020111653

