## Supplemental Materials for "Seroresponse to SARS-CoV-2 vaccines among maintenance dialysis patients over six months"

**Supplemental Methods**

The SARS-CoV-2 vaccine monitoring protocol automatically deactivated after two consecutive months with SAb-IgG titer less than 1 U/L. However, to avoid thereby selecting for patients with better seroresponse, subsequent monthly SAb-IgG levels among patients with consecutive months with SAb-IgG titer below 1 U/L were adjudicated as 0 U/L if the patient had not died, developed COVID-19, received transplantation, or transferred out of the DCI system. This adjudication is consistent with the assumption behind the protocol's deactivation, specifically that patients who do not have seroimmunity for two consecutive months are unlikely to develop or re-develop it thereafter without exposure to SARS-CoV-2 or undocumented additional vaccine dose. 215 test results were adjudicated this way.

In addition, if a patient had more than one titer assessed in a calendar month, only the first measurement was retained. Titers at the assay's upper limit of detection, 20 U/L, were coded as values of 20 U/L. Lab results were removed from the data set if they were captured after dates of transplantation or COVID-19 diagnosis; 1 and 7 such samples were excluded from analyses, respectively.

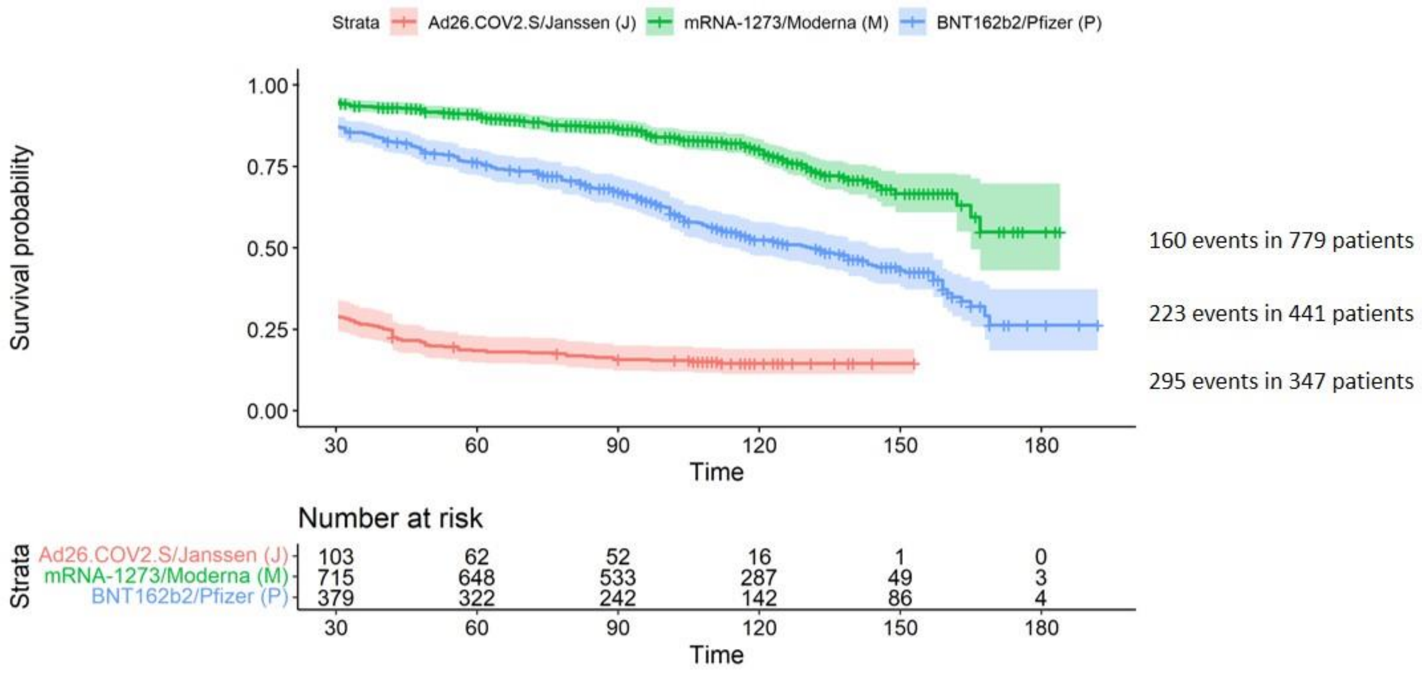


**Figure S1: Kaplan-Meier survival curves for the outcome of SAb-IgG titer <2 or diagnosis of COVID-19, among those without a history of COVID-19, by vaccine type.** Data are shown beginning at Day 30, at which time all patients have had at least one opportunity for assessment of the outcome of SAb-IgG titer < 2 U/L via monthly labs. Patients were censored at death, transplantation, or last available titer assessment.

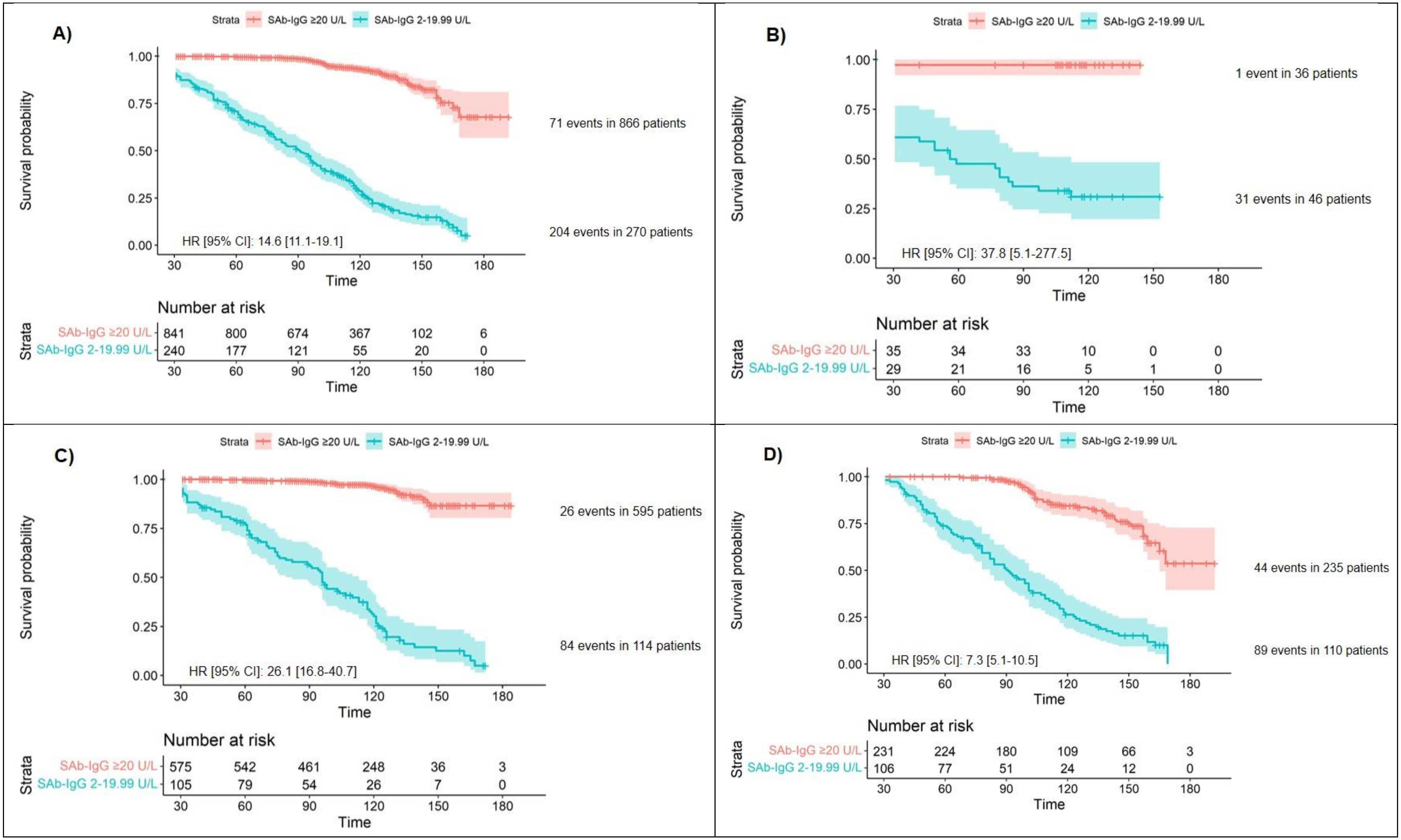


**Figure S2: Kaplan-Meier survival curves for the outcome of SAb-IgG titer <2 or diagnosis of COVID-19, among those without a history of COVID-19, by maximum initial SAb-IgG titer.** Data are shown beginning at Day 30, at which time all patients have had at least one opportunity for assessment of the outcome of SAb-IgG titer < 2 U/L via monthly labs. Patients were censored at death, transplantation, or last available titer assessment. Patients with maximum initial SAb-IgG titer are not shown since, given our definition of maximum initial titer, all had experienced the outcome by the end of month 2.

A) All patients
B) Ad26.COV2.S/Janssen recipients only
C) mRNA-1273/Moderna recipients only
D) BNT162b2/Pfizer recipients only

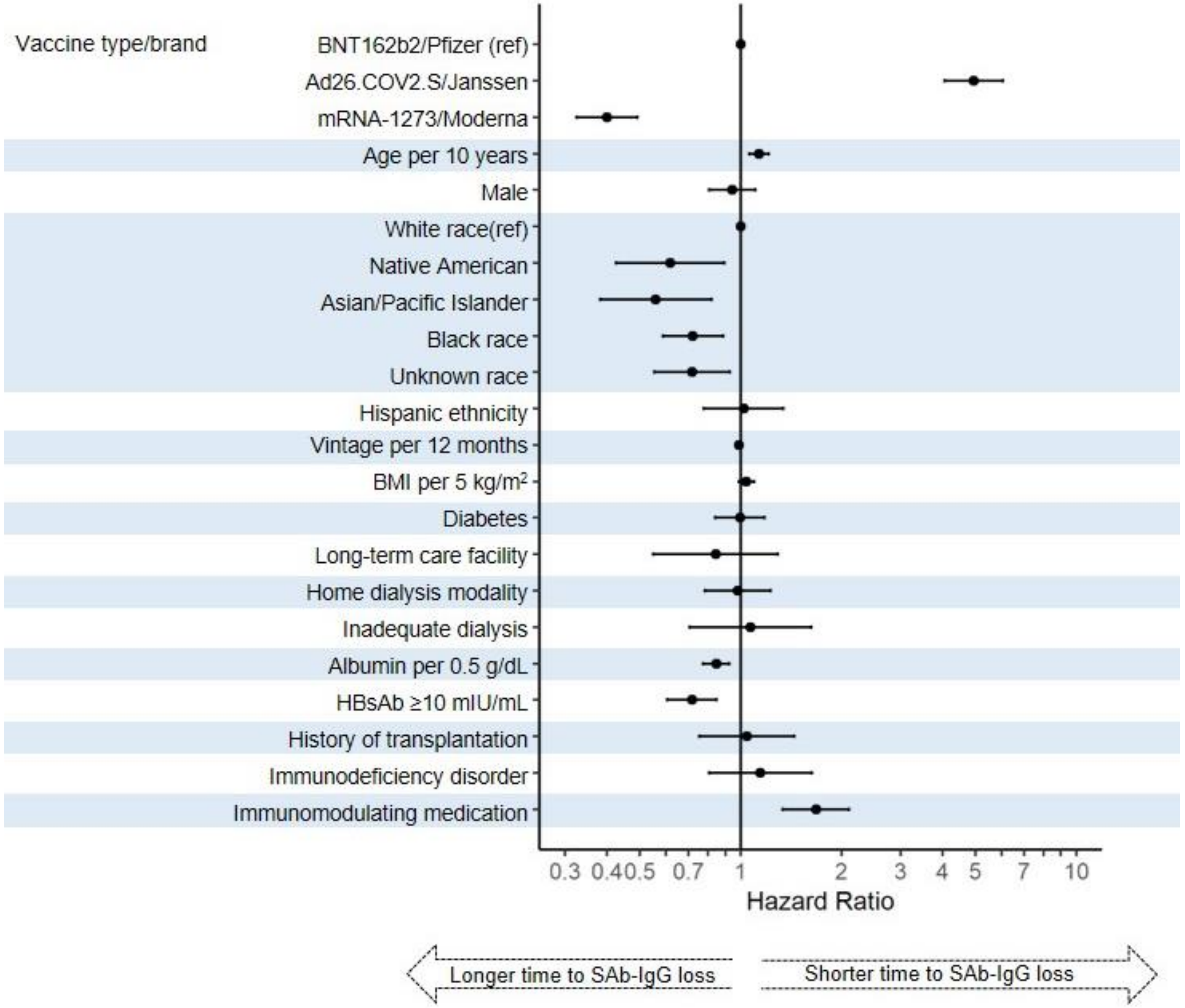


**Figure S3**. Multivariable Cox proportional hazards regression of clinical characteristics predicting loss of SAb-IgG seroresponse (outcome of SAb-IgG titer < 2 or development of COVID-19).

Inadequate dialysis defined by hemodialysis dose spKt/V<1.2 or peritoneal dialysis dose weekly Kt/V<1.7

HBsAb ≥10 mIU/mL signifies hepatitis B seroimmunity
